# Peripheral monocyte transcriptomics associated with immune checkpoint blockade outcomes in metastatic melanoma

**DOI:** 10.1101/2024.01.25.24301653

**Authors:** Rosalin A Cooper, Chelsea A Taylor, Robert A Watson, Orion Tong, Isar Nassiri, Piyush Kumar Sharma, Martin Little, Weiyu Ye, Surya Koturan, Sara Danielli, Mark Middleton, Benjamin P Fairfax

**Affiliations:** MRC Weatherall Institute of Molecular Medicine, University of Oxford, Oxford, UK; Department of Oncology, University of Oxford, Oxford, UK; Department of Cellular Pathology, John Radcliffe Hospital, Oxford University Hospitals, Oxford, UK; Cancer and Haematology Centre, Oxford University Hospitals, Oxford, UK

## Abstract

Clinical responses to immune checkpoint blockade (ICB) for metastatic melanoma (MM) are variable, with patients frequently developing immune related adverse events (irAEs). The role played by myeloid populations in modulating responses to ICB remains poorly defined. We explored the effect of MM and the response to ICB across a cohort of patients with MM (n=116) and healthy donors (n=45) using bulk and single cell RNA-seq, and flow cytometry. Monocytes from patients with MM exhibit highly dysregulated baseline transcriptional profiles, whilst ICB treatment elicits induction of interferon signaling, MHC class II antigen presentation and CXCR3 ligand expression. Although both combination (cICB - anti-PD-1 and anti-CTLA) and single-agent (sICB - anti-PD1) ICB therapy modulates a shared set of genes, cICB displays a markedly greater magnitude of transcriptional effect. Notably, we find increased baseline monocyte counts correlate with a monocyte proliferation signature and risk of early death, whilst a gene-signature corresponding to a subset of platelet-binding classical monocytes conversely associates with improved outcome. This work demonstrates a central role for monocytes in the modulation of treatment response to ICB, providing insights into inter-individual variation in immune responses to ICB and further highlighting the multifarious immunological consequences of ICB treatment.

## Main text

Immune checkpoint blockade (ICB) with single-agent anti-PD1 (sICB) or combination anti PD-1/anti-CTLA4 (cICB) monoclonal antibodies has transformed the clinical outlook for patients with metastatic melanoma (MM)^1^. However, clinical responses are variable and many patients develop immune-related adverse effects (irAEs)^2,3^. Whilst the relationship between peripheral CD8^+^ T cell characteristics and clinical response of MM to ICB therapy is well described^4,5^, myeloid responses to ICB remain poorly defined. Existing work suggests that the circulating monocyte population can modulate response to ICB, with an expanded monocyte population typically associated with impaired clinical response to anti-PD1 therapy in both MM^6^ and non-small cell lung cancer (NSCLC)^7^. Monocytes are conventionally categorised into three subsets: CD14^+^CD16^-^ classical monocytes, CD14^+^CD16^+^ intermediate monocytes and CD14^dim^CD16^+^ non-classical monocytes^8^. In MM, small cohort studies have described relationships between monocyte subset counts to response to both anti-PD1 and anti-CTLA4 treatment^9,10^. There has been limited analysis of the impact of standard-of-care combination anti-CTLA-4/anti-PD-1 treatment however, whilst the impact of ICB on monocyte gene expression and relationships with clinical outcomes are similarly unexplored.

We hypothesised that transcriptomic analysis of peripheral monocytes may provide prognostic information and novel insights ICB-treatment response in patients with MM. We assessed monocyte responses to ICB in isolated cells using bulk RNA-sequencing (RNA-seq) across 116 individuals from a cohort of MM patients receiving ICB as well as samples from 45 healthy controls. We find MM is associated with distinct pro-inflammatory changes in monocyte gene-expression, the extent of these correlating with baseline monocyte count and having prognostic value. Treatment with either cICB or sICB therapies evokes shared patterns of gene expression but no consistent change in counts of conventional subsets. We further characterised heterogeneity in peripheral monocyte subsets in MM with scRNA-seq and flow cytometry, noting relationships with clinical outcome, observing positive outcomes to be associated with an expanded platelet binding subset whereas monocyte proliferation is associated with risk of early death.

## RESULTS

### Circulating monocytes in patients with metastatic melanoma have a distinct transcriptomic profile

To identify MM-associated signatures in circulating monocytes at baseline, we performed differential gene expression (DGE) across monocytes from patients with MM versus healthy donors (HD) (C1 n = 114 samples; HD n = 45 samples). This identified 1,774 significantly differentially regulated transcripts in patient derived monocytes (P_adj_ <0.05; Figure 1A, Supplementary Table 1), including 871 upregulated, and 903 suppressed. Of note, genes encoding the chemokine receptors CXCR1 and CXCR2, which bind to pro-inflammatory cytokine and neutrophil chemoattractant IL-8, implicating pathways previously found to be associated with a less favorable response to ICB^12^, were markedly upregulated in MM patients (Figure 1B). Interrogation of the Gene Ontology Biological Processes (GOBP) database^13^ identified key expression pathways enriched in patient samples including chemotaxis (GO:0006935, P_adj_ = 0.0014; GO:0060326, P_adj_ = 0.0096), neutrophil degranulation (GO:0043312, P_adj_ = 5.4×10^-14^), response to lipopolysaccharide (GO:0032496, P_adj_ = 0.00027), and vascular endothelial growth factor (VEGF) signaling (GO:0048010, P_adj_ = 0.02) (Figure 1C, Supplementary Table 2). Given increasing monocyte count is associated with poorer clinical outcome in melanoma^6^ and non-small cell lung cancer (NSCLC)^7^, we examined the relationship between baseline monocyte gene expression and pre-treatment hospital measured monocyte count. Strikingly, increasing monocyte count was associated with markedly divergent expression profile, with 1,344 transcripts associated with pre-treatment monocyte count (88 C1 samples, P_adj_<0.05; Figure 1D, Supplementary Table 3), of which 694 transcripts were positively associated with count, the most significant being *FAM20A*, encoding a pseudokinase with putative roles in haematopoiesis^14^ (Figure 1E). Conversely, increasing counts were negatively associated with 650 transcripts, most significantly *IMPDH2*, encoding a gene involved in purine metabolism in response to hematopoietic stress^15^ (Figure 1E). Pathways positively correlated with monocyte count included neutrophil degranulation (GO:0043312, P_adj_ = 4.6×10^-20^), as well as heterotypic cell-cell adhesion (GO:0034113, P_adj_ = 2.5×10^-3^) and VEGF signaling (GO:0048010, P_adj_ = 2.9×10^-3^), whilst suppressed pathways were mainly involved in rRNA processing (GO:0006364, P_adj_ = 5.7×10^-23^) and DNA replication (GO:0006260, P_adj_ = 7.0×10^-8^) (Supplementary Table 4). Of particular significance to cancer immunology was the negative association of IFNG regulated MHC class II gene expression, crucial in antigen presentation, including *HLA-DRA* (P_adj_ = 0.0005), *HLA-DMA* (P_adj_ = 0.0063), *HLA-DPA1* (P_adj_ = 0.00057) and *HLA-DPB1* (P_adj_ = 0.0010) with peripheral monocyte count (Supplementary Table 3). In keeping with this, transcripts anti-correlated with monocyte count were enriched for pathways including antigen presentation via MHC class II (GO:0019886, P_adj_ = 0.014) and T cell co-stimulation (GO:0031295, P_adj_ = 0.041) (Supplementary Table 4). Thus, an expanding circulating monocyte compartment is characterized by polarisation towards an immature, mitotically active ‘classical monocyte’ phenotype with predisposition towards heterotypic cell adhesion, VEGF mediated angiogenesis and reduced antigen presentation function.

**Figure 1:**
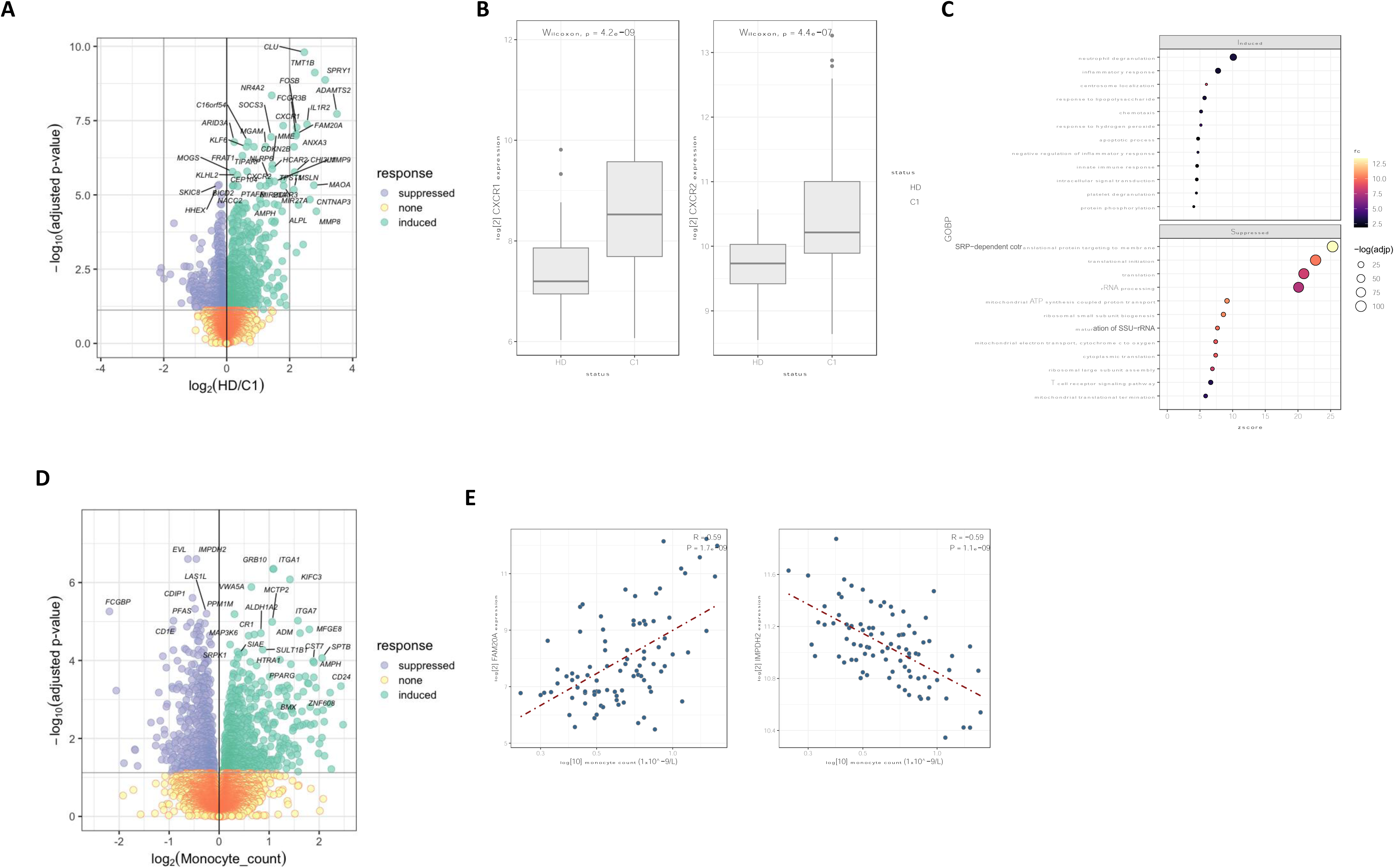
Peripheral monocytes exhibit distinct transcriptomic profiles in patients with metastatic melanoma. 1A)Volcano plot to show significantly differentially expressed genes between MM patients at baseline (‘C1’) and healthy donors (HD) (114 C1 samples, 45 healthy controls, DESEq2 analysis controlling for age and sex (P_adj_ <0.05). B) Boxplot to show log2(CXCR1) and log2(CXCR2) expression in monocytes in MM patients at baseline (C1) versus healthy donors (HD). C) Dot plot to show key enriched and suppressed GOBP pathways in monocytes in MM patients versus healthy controls (P_adj_ <0.05). D) Volcano plot to show monocyte expressed transcripts associated with baseline peripheral monocyte count (DESEq2 analysis, controlling for age and sex, 88 C1 samples, P_adj_<0.05). E) Scatterplot to show correlation between monocyte expression of FAM202 and IMPDH2 and peripheral monocyte count at baseline (C1) (Pearson’s correlation).

### ICB treatment modulates circulating monocyte gene expression

To address the impact of ICB on monocytes, differential expression analysis between baseline (pre-treatment, ‘C1’) and post-treatment (‘C2’) samples (n = 96 patients), controlling for age, sex and treatment type (sICB/cICB) was performed in bulk monocyte RNA-seq data. This identified a total of 1,067 significantly differentially modulated transcripts with ICB (758 induced, and 309 downregulated; P_adj_< 0.05) (Figure 2A & 2C, Supplementary Table 5). ICB treatment robustly induced type I and II interferon signaling (Figure 2A, Supplementary Table 5) including JAK-STAT pathway members *STAT1* (P_adj_ = 2.0×10^-7^), *STAT2* (P_adj_ = 3.0×10^-4^), and *JAK2* (P_adj_ = 1.4×10^-6^). The CXCL3 ligands *CXCL9* (P_adj_ = 8.1×10^-9^), *CXCL10* (P_adj_ = 1.0×10^-8^) and *CXCL11* (P_adj_ = 4.1×10^-6^) were similarly induced. These IFN-γ induced transcripts are involved in CXCR3-dependent immune cell chemotaxis and T_H_1 polarisation^16,17^ and are associated with development of irAEs^18,19^. Similarly induced were transcripts encoding the classical complement proteins *C1QA* (P_adj_ = 3.6×10^-5^), *C1QB* (P_adj_ = 9.8×10^-5^) and *C1QC* (P_adj_ = 4.4×10^-5^); MHC class II molecules including *HLA-DRA* (P_adj_ = 1.6×10^-4^) and *HLA-DPB1* (P_adj_ = 1.9×10^-5^), as was *CD274* (P_adj_ = 2.8×10^-9^), encoding PD-L1^20,21^ (Figure 2A, Supplementary Table 5). Correspondingly, ICB induced transcripts were enriched across multiple pathways including IFN-γ-signaling (GO:0060333, P_adj_ = 2.5×10^-21^), type I interferon-mediated signaling (GO:0060337, P_adj_ = 9.8×10^-12^), antigen presentation via MHC class I (GO:0002479, Padj = 2.5×10^-21^) and class II (GO:0019886, P_adj_= 0.0069), and T cell co-stimulation (GO:0031295, P_adj_ = 1.5×10^-4^) (Figure 2D, Supplementary Table 6). Expression of signaling pathways including tumour necrosis factor (TNF) (GO:0033209, P_adj_ = 2.5×10^-10^) and NFκB signaling (GO:0043123, P_adj_ = 5.3×10^-3^) were also enriched in monocytes following ICB (Figure 2D, Supplementary Table 6).

**Figure 2:**
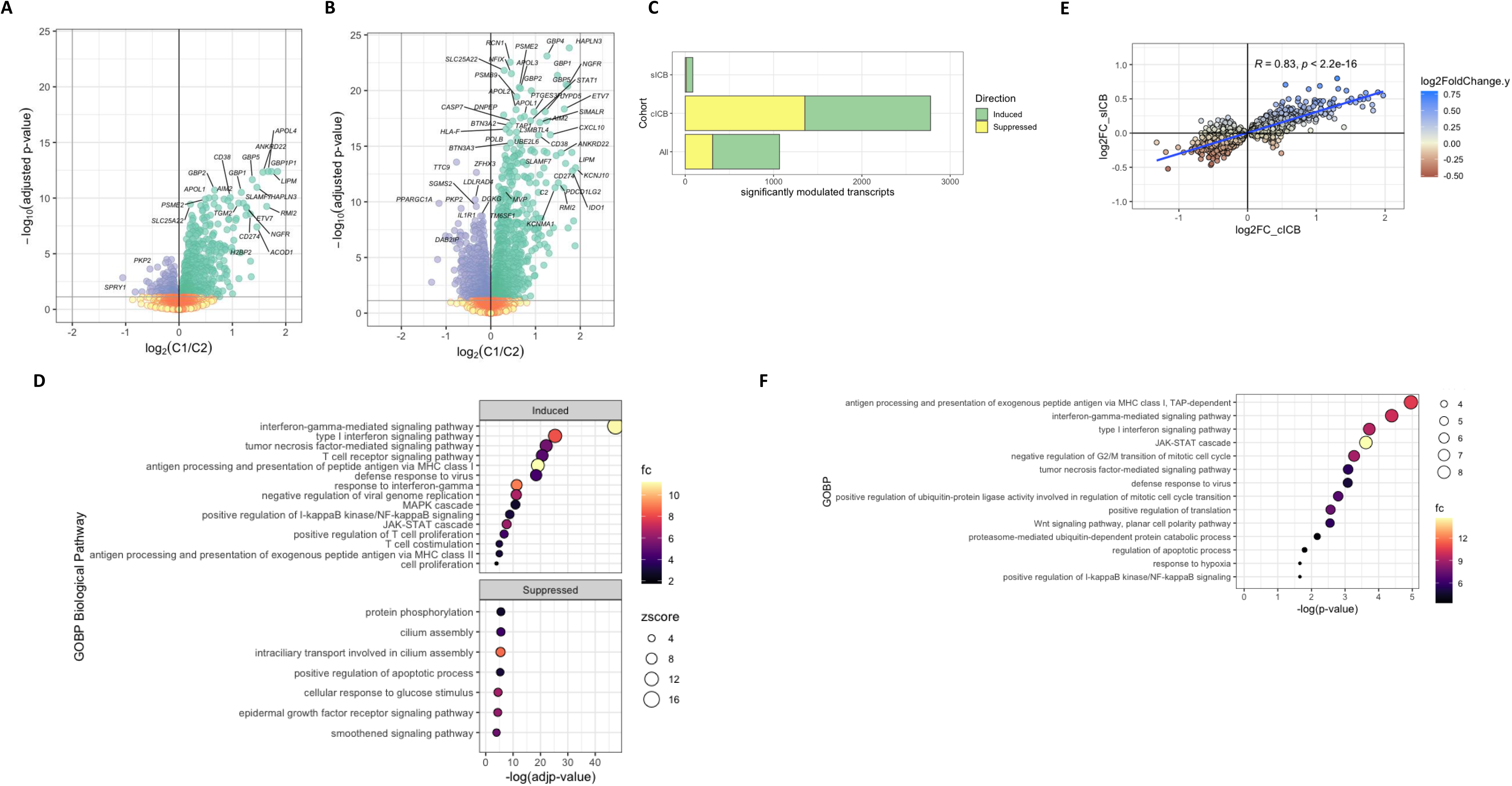
Transcriptional responses to ICB in circulating monocytes. Volcano plot to show differentially regulated transcripts in response to: A) ICB across cICB and sICB treated patients (DESEq2 controlling for age, sex and treatment type (sICB or cICB), n = 96 patients, P_adj_<0.05) and B) cICB (DESEq2 pairwise analysis, 51 patients). C) Barplot to show number of induced/suppressed transcripts with ICB (Both sICB and cICB treated patients, n = 96 patients, sICB n=45 patients and cICB n= 51 patients). D) Dot plots to show key significantly enriched GOBP pathways induced and suppressed with ICB across all patients (P_adj_<0.05). E) Plot to show correlation between log_2_ fold change (FC) of transcripts significantly differentially expressed with cICB (‘log2FC_cICB’) versus the log_2_ fold change of those same transcripts following sICB (‘log2FC_sICB’) (Pearson’s correlation). F) Dot plot to show key preferentially enriched GOBP pathways with cICB versus sICB (P_adj_<0.05).

Comparative analysis of monocyte responses to cICB versus sICB demonstrated that, as per CD8 T cells^4^, cICB was associated with a greater magnitude of transcriptional modulation, with 2,777 transcripts differentially expressed with cICB (pairwise analysis, n=51 patients) but only 85 following sICB (pairwise analysis, n=45 patients) (Figure 2B, 2C, Supplementary Figure 1, Supplementary Tables 7 & 8). There was near complete concordance of the transcripts modulated by cICB versus sICB (80/85 transcripts) and sICB and cICB demonstrated consistent shared direction of effect, but greater magnitude of transcriptional modulation with cICB (Figure 2C, 2E). A total of 352 transcripts (240 induced, 112 suppressed) were preferentially regulated in monocytes with cICB compared to sICB. Preferentially cICB induced transcripts included *CD274* (P_adj_ = 0.0033), *CXCL10* (P_adj_ = 6.3×10^-3^), *CXCL11* (P_adj_ = 0.042), and *C1QC* (P_adj_ = 0.019) (Supplementary Table 9) and corresponding pathway analysis of these genes again highlighted enrichment of type I IFN and IFN-γ-related signaling, JAK-STAT and T cell receptor signaling (Figure 2F, Supplementary Table 10). This suggests that, as with CD8^+^ T cells^4^, cICB is associated with a qualitatively similar but quantitively greater transcriptional effect on peripheral monocytes.

### Peripheral monocyte transcriptomic profiles predict clinical outcome

We then identified monocyte-expressed markers of clinical response by performing differential expression analysis of bulk RNA-seq data with clinical outcome parameters. Clinical parameters including risk of death at three, six and twelve months after date of first treatment, and progression at six-months were used to dichotomise patients, with differential expression analysis controlled for age, sex and, where applicable, treatment type and cycle. We found a monocyte cycling signature characterised by expression of proliferation index marker *MKI67* to be associated with death at all three time points (Figure 3A-C, Supplementary Tables 11-13) with 314, 239 and 444 transcripts associated with death at three, six and twelve months respectively (controlling for age, sex, treatment status and type across 114 C1 and 98 C2 patient samples, P_adj_<0.05) (Figure 3A-C, Supplementary Tables 11-13). Transcripts across all time points positively correlated with death were enriched for pathways associated with the monocyte MM-associated expression signature we describe above, including neutrophil degranulation (GO:0043312), platelet degranulation (GO:0002576) and response to LPS (GO:0032496) (Figure 3E, Supplementary Table 14-16). Strikingly, when we explored this by treatment status, risk of death was primarily related to pre-treatment samples (Supplementary Figure 2, Supplementary Table 17A-C). We also found 269 transcripts associated with progression at six months (113 C1 samples and 98 C2 controlling for treatment type, status, age and sex, P_adj_<0.05) (Figure 3D, Supplementary Table 18A) which were enrichment for pathways including angiogenesis (GO:0001525) (Supplementary Table 18B). These observations indicate previously undescribed mechanisms through which the prognostic effects of peripheral monocytes and elevated counts are mediated.

**Figure 3:**
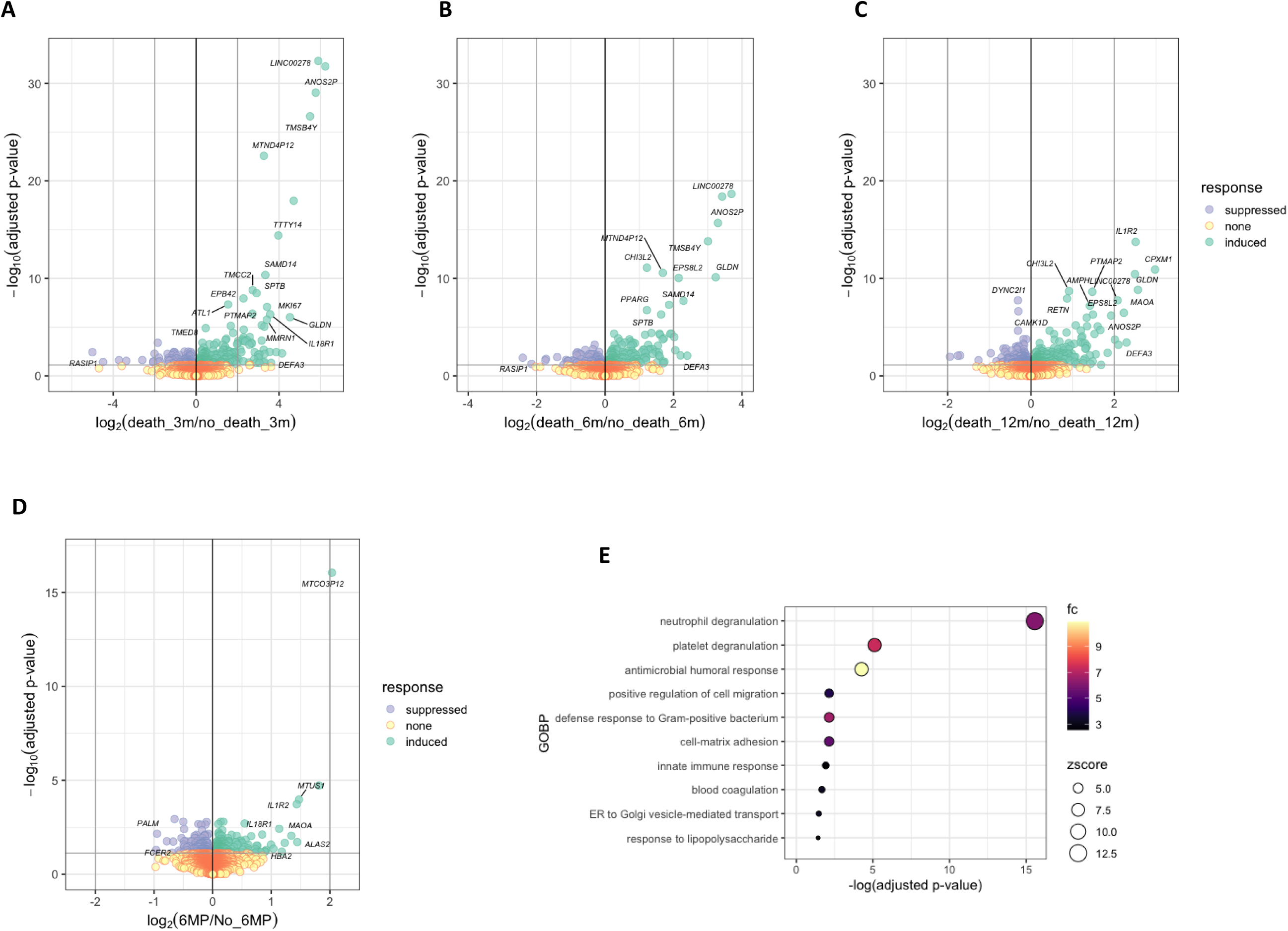
Transcriptional correlates of clinical response in peripheral monocytes. Volcano plot to show transcripts associated with death at: A) three months; B) six months and C) twelve months after the date of commencing of the first cycle of ICB treatment (DESeq2 analysis, controlling for age, sex, treatment status and treatment type, 114 C1 patients, 98 C2 patients, P_adj_ <0.05). D) Volcano plot to show transcripts associated with disease progression at six months (DESeq2 analysis, controlling for age, sex, treatment status and treatment type, 113 C1 patients, 98 C2 patients, P_adj_ <0.05). E) Dot plot to show selected GOBP pathways enriched across transcripts associated with risk of death at three months (P_adj_<0.05).

### Dissecting heterogeneity in monocyte subgroup responses to ICB

Monocytes are heterogeneous in composition but are conventionally categorized into three subsets: CD14^+^CD16^-^ classical monocytes, exhibiting prominent chemotactic properties, CD14^dim^CD16^+^ non-classical monocytes implicated in antibody dependent cellular cytotoxicity (ADCC)^23^; and CD14^+^CD16^+^ intermediate monocytes, characterized by prominent MHC class II expression^24^. We used flow-cytometry to explore monocyte populations pre and post ICB, finding no significant change in any subset size with treatment (n = 53 paired samples) (Figure 4A), this observation remaining when performing sICB and cICB specific analysis (Supplementary Figure 3). Notably, survival analysis showed that a larger proportion of cytometry-identified non-classical monocytes at both baseline and following treatment was associated with prolonged overall survival (OS) (at baseline C1 P = 0.0079; post-treatment C2, P = 0.0016, log-rank tests) and extended progression free survival (PFS) (P = 0.014 at C1, post-treatment C2: P = 0.02, log-rank test) (Figure 4B). Conversely, consistent with the hospital blood monocyte count, increasing monocyte proportion as a total of peripheral blood mononuclear cells (PBMCs) pre-treatment was associated with reduced OS (pre-treatment, P= 0.014, log-rank test), and post-ICB reduced PFS and OS (post-treatment PFS P = 0.028; OS, P= 0.031, log rank test) (Figure 4C). In keeping with previous studies^25^, a higher lymphocyte to monocyte ratio (LMR) at baseline was associated with extended OS (p = 0.035, log-rank) (Supplementary Figure 3).

**Figure 4:**
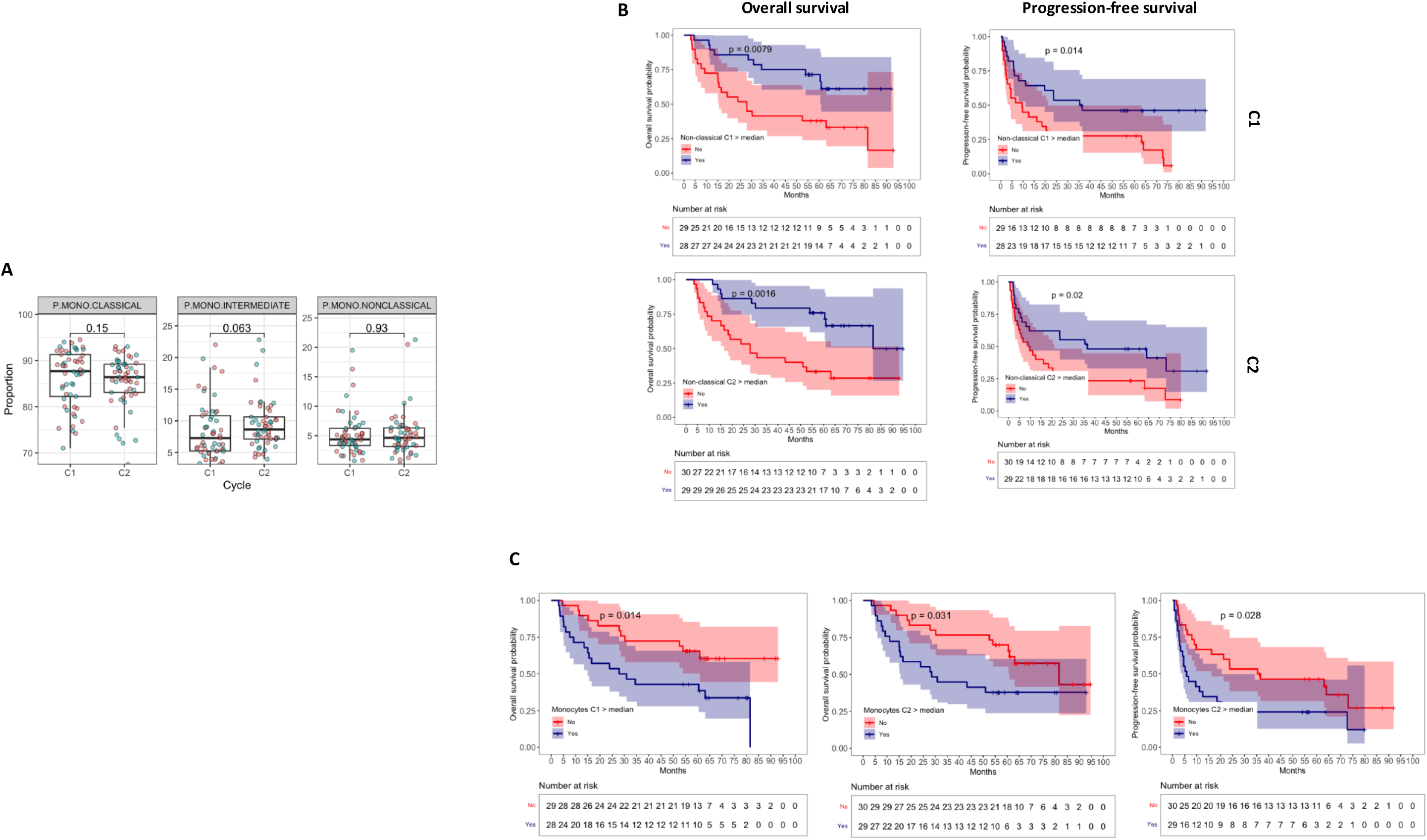
Monocyte heterogeneity and immune checkpoint blockade responses. A)Boxplots to show monocyte subsets (classical, intermediate and non-classical) as a proportion of the total peripheral monocyte population as assessed by flow cytometry across 53 paired C1 and C2 patient samples (paired Wilcoxon signed-rank test). Points are coloured by treatment type (cICB = red, sICB = blue). B) Kaplan-Meier curve to show non-classical monocyte subset size as a proportion of monocyte population above/below median) and overall survival (OS) (C1, n=57, C2, n=59) and progression free survival (PFS) (57 C1, 59 C2 samples) (log-rank test). C) Kaplan-Meier curve to show total monocyte subset size as a proportion of all PBMCs on flow cytometry above/below median) and OS at baseline at C1 (n = 57 patients, log-rank tests), and C2 (n=59 patients, log-rank test) and progression free survival (PFS) at C2 (n = 59, log-rank test).

### Characterising peripheral monocytes with scRNA-seq

To further characterize the predictive role of monocyte subsets, we performed scRNA-seq of monocytes from peripheral blood both pre- and post-ICB treatment from eight patients with MM, as well as three healthy donors (Supplementary Figure 4). Unsupervised clustering of cells across all conditions was performed to a high resolution i.e. ‘overclustering’, and comparison with publicly available annotated expression datasets using *SingleR* (Supplementary Figure 4) was performed. Monocyte groups present across all conditions/patients were selected for downstream analysis (Supplementary Figure 5) and a total of 22,116 cells were analysed. Two main monocyte subsets were identified: those aligned to a classical monocyte annotation, and those annotated as an intermediate/non-classical *FCGR3A*-expressing ‘CD16^+^ monocyte’ group (Figure 5A, Supplementary Figure 5). The large classical-like monocyte cluster was characterised by markers including *CD14* (P_adj_= 5.3×10^-251^), and genes involved in adhesion and migration including *CLEC4E*^29^ (P_adj_= 6.6×10^-67^), *VCAM*^30^ (P_adj_ < 1×10^-300^), *CCR2*^29,31^ (P_adj_= 2.0×10^-66^) and *SELL*^29^ (P_adj._ = 1.0×10^-94^) (Supplementary Figure 6, Supplementary Table 20). Pathway enrichment included response to lipopolysaccharide (GO:0032496) and response to wounding and wound healing (GO:0009611, GO:0042060) (Supplementary Figure 6, Supplementary Table 22). CD16+ monocytes were defined by expression of *FCGR3A* (P_adj_ < 1×10^-300^), *TNF* (P_adj_= 6.2×10^-52^), classical complement genes *C1QA* (P_adj_= 2.2×10^-167^)*, C1QB* (P_adj_= 1.51qx10^-116^), and *C1QC* (P_adj_= 4.7×10^-83^) and interferon-induced genes including *IFITM1* (P_adj_= 5.8×10^-181^)*, IFIT2* (P_adj_= 2.2×10^-22^), and *IFIT3* (P_adj_= 7.3×10^-73^) (Supplementary Table 20), these genes being enriched for T cell co-stimulation (GO:0031295), T cell receptor signaling (GO:0050852), and antigen processing and presentation via MHC-class II pathways (GO:0019886) (Supplementary Table 22) pathways.

**Figure 5:**
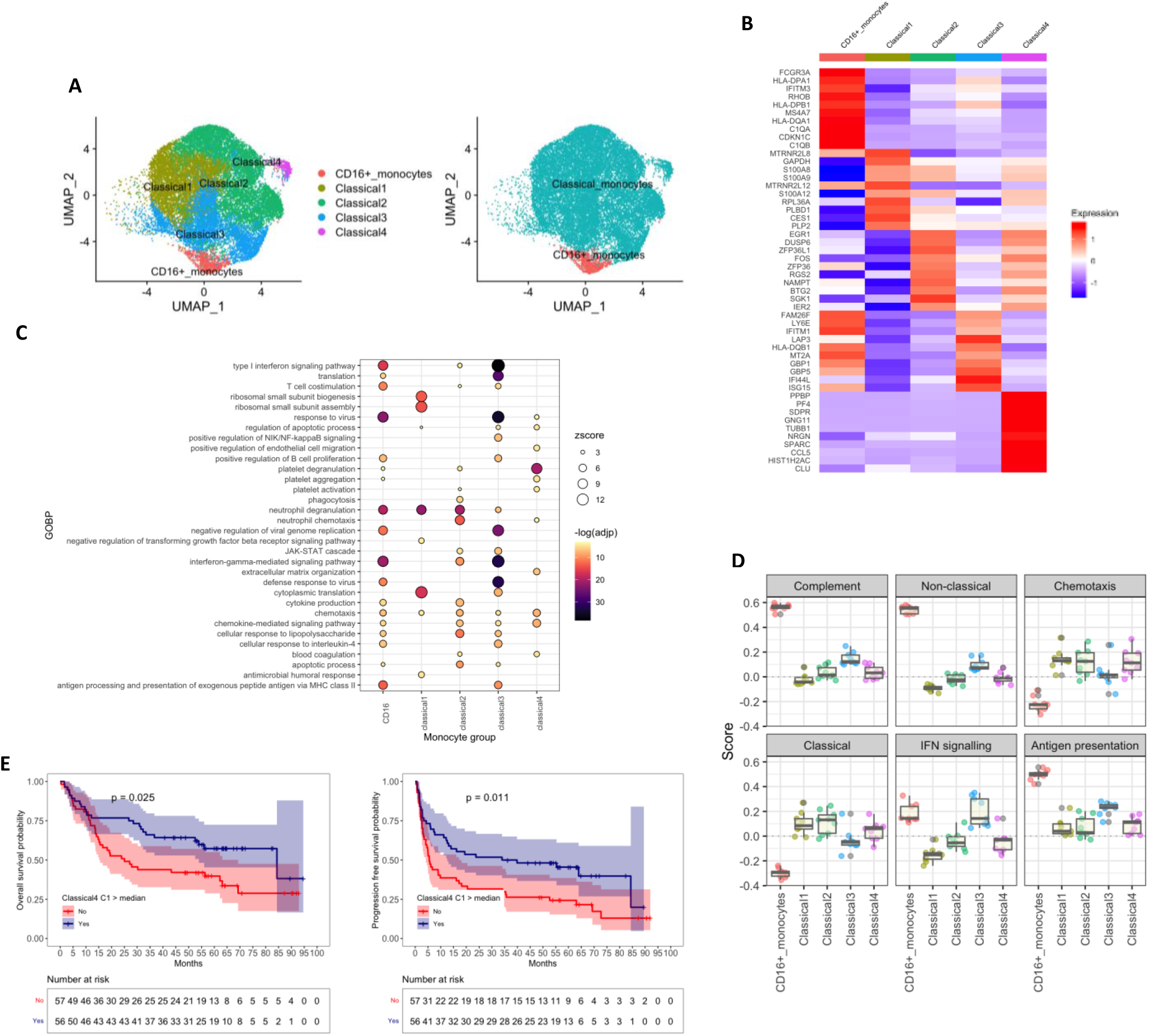
Characterising monocyte subsets in the context of immune checkpoint blockade. A) (Left) UMAP plot to show monocytes from eight patients receiving ICB at baseline (C1, d0) and at d21 post-treatment (C2), plus three healthy donors) and Classical clusters 1-4 and CD16+ monocytes (total cells n=22,116). (Right) UMAP plot to show monocytes coloured according to classical and CD16^+^ cluster annotation assignment. B) Expression heatmap showing gene expression profiles for Classical1, Classical2, Classical3, Classical4 and CD16^+^ monocytes groups. Genes were selected to include the ten genes per classical group with the greatest log2 fold change. C) Dotplot to show key GOBP pathways enriched in Classical1-4 and CD16^+^ monocyte groups. D) Gene expression module score for each monocyte subgroup. Score is average of expression of top 50 subgroup-defining genes for each monocyte subgroup. Positive value infers expression is higher than expected, and negative value lower. F) Kaplan-Meier curve to show Classical4 expression score in bulk RNA-seq data above/below median expression score) at C1 and OS (n = 113 patients, log-rank test) and PFS (n = 113 patients, log-rank test).

We did not identify a distinct intermediate monocyte population on high-level unsupervised clustering, however unsupervised clustering of the large classical monocyte compartment revealed further heterogeneity within it. We found the classical monocyte compartment could be further divided into four distinct sub-clusters: ‘Classical1’ characterised by high expression of alarmin complex genes *S100A8/A9* ; ‘Classical2’ with high expression of *CD14* and *EGR1,* a key inhibitory regulator of myeloid populations^28^; ‘Classical3’ with robust expression of an IFN-response associated signature including *GBP1, GBP2,* and *STAT1*; and ‘Classical4’ which had a distinct monocyte-platelet aggregate (MPA)-like classical profile with high expression of platelet-associated genes including *PPBP* (P_adj_ = 9.5×10^-159^) which encodes chemokine and CXCR1/CXCR2 ligand CXCL7 in activated platelets^26^, and *GP9* (P_adj_ = 2.3×10^-38^), a von Willebrand factor receptor required for clotting^27^ (Figure 5A & 5B, Supplementary Table 19). Pathway enrichment analysis of group-defining markers demonstrated overlap between groups, but Classical1 was characterised by enrichment of antimicrobial humoral response pathway, Classical2 for phagocytosis, Classical3 for defense response to virus, antigen processing via MHC class II and T cell co-stimulation (Figure 5C, Supplementary Table 21) whilst Classical4 was enriched for platelet activation and blood coagulation pathways (Figure 5C, Supplementary Table 21). Notably, although power to detect ICB related subset size changes was limited, we found no significant change in the proportion of classical, CD16^+^ or classical subgroups (Classical_1_ – Classical_4_) with ICB (Supplementary Figure 7), in keeping with flow-cytometry observations.

We performed analysis of monocytes according to expression of gene sets consisting of the most significantly associated transcripts of a candidate ‘hub gene’ (Supplementary Figure 8). Both antigen-presentation and complement module scores comprised genes characteristic of non-classical monocytes such as *FCGR3A, CDKN1C* and MHC Class II genes (Supplementary Tables 23-28). The chemotaxis score comprised a classical expression signature. CD16^+^ monocytes had higher expression antigen-presentation, complement production, and IFN signaling modules, whilst classical monocytes exhibited higher expression of chemotaxis-associated gene modules (Figure 5D). Expression module scoring revealed a continuum of activation states across the classical subgroups, with Classical1 and Classical2 exhibiting low complement and antigen presenting expression module scores, and Classical3 expressing higher expression of these modules (Figure 5D).

Having defined monocyte transcriptional heterogeneity with scRNA-seq in a subset of patients, we determined to explore the clinical correlates of the identified subset associated expression signatures across the full cohort. Intriguingly, we found that expression above the median of the MPA expression signature (Classical4) at baseline was associated with improved clinical outcomes to ICB with significant associations to PFS (p = 0.011, log-rank median PFS low MPA: 5.5 mnths (95% CI 15.2 – 30.2), median PFS high MPA: 35.1 mnths (95% CI 28.1 – 43.1) and OS (P = 0.025, log-rank, mean OS low MPA: 25.0 mnths (95% CI 28.1 – 43.1), median OS high MPA: 53.3 mnths (95% CI 38.4 – 52.3)) (Figure 5E).

## DISCUSSION

Cancer has been associated with altered transcriptional profiles in monocytes with upregulation of chemotaxis pathways^32–35^, impaired cytotoxicity^36^, polarisation towards an immature phenotype^37^ and altered metabolic pathways^9,32^. Here we show in the analysis of a large patient cohort that MM likewise has distinct effects on monocyte expression profiles, with induction of chemotaxis, adhesion and angiogenesis-associated pathways. An elevated monocyte count has been associated with poor prognosis across cancer types^38–41^. For the first time we show that a raised monocyte count is accompanied by profound changes in monocyte transcriptional profiles, indicating mechanisms through which the prognostic effects of elevated monocytosis may be mediated. Specifically, as well as increased expression of mitotic factors, there is upregulated VEGF expression, indicating the emergence of a cycling, immature pro-angiogenic ‘classical-type’ expression profile.

We provide novel insights into the effects of ICB treatment on immunity. Using bulk monocyte RNA-seq data from a large MM cohort we demonstrate that ICB promotes a monocyte transcriptional profile in keeping with a T cell costimulatory phenotype, with upregulation of antigen presentation via MHC class II pathways, classical complement and pro-inflammatory cytokines. Notably, *CXCL9, CXCL10* and *CXCL11* are all robustly upregulated in monocytes by ICB. The CXCL9-11/CXCR3 axis is implicated in immune migration and the development of Th1 cells^17^ whilst the myeloid CXCR3-CXCL9/10/11 axis has a key role in modulating CD8^+^ T cell responses in the context of ICB^42^. Induction of this axis in macrophages following cICB is also described, with CXCL9-expressing macrophages required for CD8^+^ T cell infiltration and for effective clinical response^43^. Here we extend these observations from murine tumour based myeloid cells, demonstrating activation of this axis in circulating monocytes in patients, indicating that monocytes may play an early role in modulating peripheral CD8^+^ T cell recruitment. Notably, as in CD8^+^ T cells, although it is qualitatively concordant the magnitude of transcriptional response is far greater with cICB versus sICB^4^. Given the dosing of nivolumab in cICB is 1mg/kg as opposed to fixed monthly dosing of 480mg, patients typically receive much less anti-PD-1 treatment at the outset of this regimen. This marked divergence in transcriptional responses between sICB and cICB attests to a highly significant synergistic effect of the addition of ipilimumab and may reflect the greater propensity for cICB to both induce irAEs as well as long-term durable disease control. Interestingly, despite marked transcriptional changes, consistent shifts in the size of monocyte proportion or classical and non-classical populations were not seen in response to ICB in our dataset, potentially indicating a peripheral ‘steady state’ following ICB in early post-treatment timepoints.

Expansion of circulating monocytes is associated with a less favourable clinical outcome across different malignancies^44–47^. In keeping with previous reported associations, we find that a smaller monocyte population both before and after ICB is associated with better clinical outcome, as is a higher pre-treatment lymphocyte to monocyte ratio. However, intra-compartmental heterogeneity, polarization and activity also appears significant. We show that a monocyte cycling signature (inferred by *MKI67* expression) with enrichment of MM-associated ‘classical-type’ signature at baseline is predictive of early death. We also explore the role of monocyte subsets in this context. A larger non-classical monocyte subset, both at baseline and following treatment, is associated with a more favourable prognosis. This suggests that response to ICB is determined in part by baseline characteristics in circulating monocytes, with an immature classical ‘cycling’ signature a poor prognostic signature, and a non-classical population associated with more favourable response to ICB. This may be underpinned by the T cell co-stimulatory phenotype and effects of pro-inflammatory non-classical monocytes.

We use scRNA-seq to dissect the roles of monocyte subsets in predicting response to ICB. Whilst there are three conventionally recognised monocyte subsets, scRNA-seq studies are notable for the discrepancy and variety of described subsets^48–51^. However, in keeping with most studies, we identify a large classical subset, and smaller CD16+ subset. We isolated monocytes for scRNA-seq with magnetic bead sorting using CD14 antibodies, and this potentially enriched for classical monocytes, allowing us to describe the transcriptional heterogeneity within this compartment. In keeping with this, our unsupervised clustering of scRNA-seq data identifies four sub-clusters within the classical subset. Notably, we observe one, Classical4, with expression profiles consistent with an MPA-associated signature. Applying the expression signature of this subset cohort-wide across the bulk monocyte RNAseq data demonstrates that increased pre-treatment expression of this geneset is associated with a more favourable prognosis. Whether this reflects a genuine subset of increasingly reactive monocytes or alternatively, is a proxy marker for anti-tumour monocyte or platelet activity is unclear and requires analysis in larger single-cell datasets currently being generated. Notably MPA are described across multiple conditions and have been associated with poor prognosis in cardiovascular disease^52^ and COVID-19^53^. Monocyte-platelet adhesion enhances CD16 expression and promotes a pro-inflammatory phenotype in circulating monocytes^54^. In the context of cancer this monocyte ‘stickiness’ at baseline may predict propensity to polarisation towards a pro-inflammatory myeloid response with T cell costimulatory effects.

A limitation of this work is the extent to which the changes we observe in monocyte expression in response to ICB therapy reflect ICB induced T cell activation, versus direct modulation of monocytes on T cell activation. Our analyses of CD8^+^ T cells in patients with MM has provided critical insights into how T cell responses and clonal dynamics reflect clinical response to ICB and confirmed the potential role for peripheral immune signatures as clinical predictive biomarkers^4,5^. Further work to dissect this interaction in larger datasets at single-cell resolution will be key to fully understanding the mechanism of immune modulation of ICB treatment across individuals and the relationship with clinical response. Nonetheless, our analysis describes favourable associations with clinical outcome of increased counts of non-classical monocyte subsets as defined by flow-cytometry. Conversely, we find that the increased monocytosis frequently observed in MM is associated with profoundly pro-inflammatory and pro-angiogenic expression profiles of monocytes in this state, with a presumed pro-tumourogenic effect, and indicating a causal role in impaired response to ICB and early death in MM.

## MATERIALS AND METHODS

### Samples

Peripheral blood samples were obtained from patients >= 18 years old with metastatic melanoma (MM) treated with immune checkpoint blockade (ICB). This included patients treated with ipilimumab/nivolumab (anti-CTLA-4/PD-1) combination immune checkpoint blockade (cICB), and with single-agent pembrolizumab or nivolumab (anti-PD-1) immune checkpoint blockade (sICB). All patients provided prior written informed consent for samples to be donated to the Oxford Radcliffe Biobank (Approved by South Central – Oxford C Research Ethics Committee 12^th^ April 2019, REC Reference 19/SC/0173 with internal Oxford Centre for Histopathology Research ethical approval, 16/A019, 18/A064) to be used for research. Peripheral blood samples were obtained prior to the first treatment dose (cycle ‘C1’, day0), and then just before the second dose (cycle 2, C2, day21). ICB treatment regimes were according to standard treatment protocols. For patients receiving cICB this involved administration of ipilimumab and nivolumab every three weeks for up to four doses. For patients treated with sICB, pembrolizumab was administered once every three weeks, or every four weeks for nivolumab. Up to 50 ml of peripheral blood was collected per sample in ethylenediaminetetraacetic acid (EDTA) tubes. Control samples were obtained from the Oxford biobank after informed written consent was obtained. Samples were collected with local ethical approval (REC 06/Q1605/55).

### Clinical Data

Clinical outcome data across the cohort was obtained, including parameters such as progression at six months, time-to-progression, overall survival (OS), progression free survival (PFS) and death status. OS was defined as the time between the start date of ICB treatment to death, whilst PFS was defined as time between the first ICB dose and disease progression according to either death or disease progression on imaging. Clinical outcome data was available for 114 samples at baseline (C1) and 98 post-treatment (C2) samples.

### Bulk RNA Sequencing

Bulk RNA-seq was carried out on CD14^+^ cells from 45 healthy donors, and 116 MM patients treated with ICB. Patients receiving adjuvant ICB were excluded from analysis. A total of 114 C1 (day0) and 98 C2 (day21) samples were sequenced, including from 91 patients with paired pre and post treatment data. Bulk RNA-seq libraries were sequenced in three batches. The first RNA cohort was sequenced using 75 bp paired-end sequencing on an Illumina Hiseq-4000 platform. The second cohort was sequenced using the Illumina NovaSeq platform with 150 bp paired-end sequencing. The third batch of samples was run on a NovaSeq6000 platform using 150 bp paired-end sequencing. The bulk RNA-seq was performed at the Oxford Genome Centre, Wellcome Centre for Human Genetics. *HISAT2*^55^ was used to align reads in FASTQ files to the GRCh38/hg38 genome build. Adequately mapped reads were identified according to MAPQ score with *bamtools. Picard* was used to remove duplicate reads. *HTseq*^56^ was used to generate read count data.

### Differential expression analysis

Differential expression analysis of bulk RNA-seq data was carried out using *DESeq2*^11^. *DESEq2* was used to normalise read counts. The *ComBat-seq*^57^ package was used to remove the effect of batch sequencing.

For differential expression analysis of transcripts expressed across healthy donors (‘HD’ or ‘C’) versus pre-treatment samples, variables including sex and sex were included in the *DESEq2* design argument (i.e. design = ∼ sex + age + cycle). Transcripts were filtered and only those with a mean count > 10 were selected for differential expression analysis. Differential expression analysis was performed using a binomial Wald test. The Benjamin-Hochberg (BH) method was used to correct for multiple testing to identify significantly differentially expressed transcripts (P_adj_<0.05).

To identify transcripts in CD14^+^ cells associated with clinical outcome parameters, differential expression analysis was used, dichotomising clinical outcome data, either into those who had disease progression at six months, versus those who did not progress; and those who died three, six and twelve months after starting ICB treatment, versus those that did not. *DESEq2* was used to perform differential expression analysis, controlling for treatment state and type (sICB or cICB) where applicable, as well as age and sex.

### Pathway enrichment analysis

The *XGR*^58^ package was used for pathway enrichment analysis using interrogation of the GOBP database. Unless otherwise stated, induced/suppressed or positively/negatively associated transcripts were analysed separately.

### scRNA-seq samples

scRNA-seq library preparation and sequencing was performed as previously described^4,5^. In the first batch, CD8^+^ and CD14^+^ cells were isolated from PBMCs from peripheral blood from four patients treated with ipilimumab/nivolumab combination-immune checkpoint blockade (cICB) and four treated with pembrolizumab single-immune checkpoint blockade (sICB). Samples were obtained before (C1, day0) and after (C2, day21) the first treatment cycle. Healthy donor samples from three individuals were pooled prior to library preparation and sequenced in a second batch.

### scRNA-Seq library preparation

The Chromium 10x platform was used for scRNA-seq library preparation. scRNA-seq data was prepared in two batches. In the first batch, 6000 of both CD8^+^ and CD14^+^ cells were combined3 in suspension (total number 12,000 cells). In the second batch, 2000 CD14^+^ cells from each of three healthy donors were mixed in suspension in a single well. 5’ transcriptome processing was performed using the Chromium 10x system. Samples were processed according to standard manufacturer’s protocols^59^. This involved initial barcoding with a unique tag, reverse transcription, cDNA amplification and library preparation.

For the first batch, 5’ transcriptome library sequencing was performed using a HiSeq platform with 75bp paired-end sequencing with a depth of at least 50,000 reads per cell. The healthy donor control samples were sequenced using a NovaSeq 6000 platform with 150 bp paired-end sequencing with a mean sequencing depth of 78,000 reads per cell. The *Cellranger* package was used to process scRNA-seq data^60^. The Cellranger *mkfastq* was used to produce FASTQ files from the Illumina BCL files. Cellranger *count* was then applied to each FASTQ file to produce a feature barcoding and gene expression library. Cellranger *mkfastq, count* and *aggr* were used to combine samples for merged downstream analysis.

### scRNA-seq quality control

For quality control (QC), the *scater* package^61^ was used to remove outliers, and *scran*^62^ package was used to remove doublets. Cells were then selected based on *CD14* expression, with those cells expressing *CD8A, CD3D, CD3E, CD3G, CD56, CD19* or *CD20* removed from downstream analysis to remove possible contaminants. QC was then performed by removing cells that had fewer than 300 different features (genes), fewer than 500 UMIs (transcripts) and those whereby more than 20% of expressed genes were mitochondrial. Genes expressed in fewer than five cells were also removed from downstream analysis.

### Clustering analysis

The *Seurat*^63^ package in R was used for downstream analysis of the scRNA-seq data. The raw gene expression matrix was normalised and then subject to *FindVariableFeatures* function^63^. Both untreated and treated samples were merged following normalisation. Integration anchors between MM patient and Healthy donor (Control) data sets were identified using *FindIntegrationAnchors* and integration of the patient and Control datasets was performed with *IntegrateData* function. Data was then scaled, and then a principal component analysis was run using the first 13 dimensions. This level of dimensionality was chosen as it was the level at which the variation between consecutive principal components was minimal as seen on an *Elbowplot*. Unsupervised clustering was performing using the *FindClusters* function and the clusters visualised using a uniform manifold approximation and projection (UMAP) plot.

The *clustree*^64^ package was used to visualise monocyte clustering trees at resolutions of 0.1, 0.2 0.25, 0.5, 0.75, 1,2 and 3 (at 13 dimensions). To perform annotation of monocyte clusters with published gene sets the *singleR*^65^ package was used to compare the scRNA-seq data to annotated expression datasets including the Human Primary Cell Atlas dataset^66^, MonacoImmune dataset^67^, Immune Cell Expression data set^68^ and scRNA-seq data published by Villani et al^48^. Only groups found across all conditions and patients were used for downstream analysis. As such, cluster 5 was removed from downstream analysis as it was present only in patient ID7 post-treatment and in controls.

### Identification of cluster-defining markers

To identify genes differentially expressed in monocyte cluster the *FindAllMarkers* function was used to find differentially expressed genes between a single subset and the remaining data, selecting only positively expressed transcripts with the default Wilcoxon signed-rank sum test. The logFC threshold was set to -Inf and a method based on Bonferoni adjustment was used to correct for multiple testing.

### Gene expression module scoring

To calculate the phenotype scores (e.g. immaturity, complement, antigen presenting scores), the Seurat *AddModuleScore* function was utilised to assign a score indicating the average expression of a defined gene set per single cell. The gene sets used for module scoring applied to each cell were identified by selecting the top 50 genes most significantly correlated with hub genes of interest such as *CD14, C1QA,* and *HLA-DR* expression. To identify significantly correlated transcripts, Pearsons correlation analysis was performed across all single cells and across Variable feature genes (n=2000) using the *Hmisc* package in R, adjusting for multiple testing using the Holm’s method with the *RcmdrMisc* function. Hub genes were selected based on association with a particular phenotype or function. For example, *FCGR3A* was selected as a hub gene indicative of non-classical phenotype.

### Subset scoring in bulk RNA-seq data

We identified the 20 most highly expressed genes identified using *FindAllMarkers* definitive of each monocyte cluster and the geometric mean score of this gene set across each *DESEq2*-normalised bulk RNA-seq patient sample was calculated. Association between this expression signature and clinical outcome data was performed using the *survminer* and *survival* packages to perform survival analysis.

### Flow cytometry

Following separation, PBMCs were stored in liquid nitrogen in 90% FCS and 10% dimethyl sulfoxide (DMSO). Samples were thawed and then 1×10^6^ PBMCs were placed into a solution of HBSS with 5% FCS on ice. After 30 mins cells were fixed in 2% formaldehyde. Samples were stained using a fixable amine-reactive viability dye (LIVE/DEAD Fixable Near-IR Dead Cell Stain Kit). The appropriate antibodies were added to the sample as outlined below. The flow cytometry was carried out with a LSRII (Becton Dickinson) platform. FlowJo software was used to analyse cells blinded to clinical outcome data. For gating, myeloid cells were identified using FSC-A and SSC-A, excluding doublets. Dead/dying cells were removed via the selection of viability stain negative cells. Monocytes were then selected based on CD14 expression, and then classical, intermediate and non-classical monocytes were gated according to relative CD14/CD16 expression.

| Antibody/stain | Fluorochrome | Clone | Dilution | Source |
| --- | --- | --- | --- | --- |
| Live/Dead cell stain kit | Near-IR | n/a | 1000 | ThermoFisher Scientific |
| CD14 | PE/Cy7 | M5E2 | 50 | BioLegend |
| CD16 | FITC | 3G8 | 50 | BioLegend |

### Survival analysis

The *survminer* and *survival* packages were used to perform survival analysis (exploring association with OS and PFS) and visualise Kaplan-Meier curves. Samples were dichotomised into two subsets according to variable of interest e.g expression score, being above or below the median value. A log-rank test was used to determine statistical significance.

### Statistical analyses

All statistical analyses were performed within the R package (v 4.2.2). Plots were created using *ggplot2.* Boxplots demonstrate the upper and lower quartiles, with the line in the middle indicating the median, and Tukey’s whiskers indicating the range. Unless otherwise states, a Wilcoxon signed-rank test was used to test for significance.

## Data Availability

All data produced in this study are available upon reasonable request to the authors and will be disseminated through online repositories at the time of publication.

---

***Supplementary Figure 1: Differentially expressed transcripts with sICB***

*Volcano plot to show differentially regulated transcripts in response to sICB (DESEq2 pairwise analysis, 45 patients)*.

***Supplementary Figure 2: Transcriptional correlates of clinical response in peripheral monocytes from patients at baseline***

*Volcano plots to show transcript expression at baseline before treatment (C1) associated with death at: A) three months; B) six months and C) twelve months after the date of commencement of first cycle of ICB treatment (DESeq2 analysis, controlling for age, sex, and treatment type, 114 C1 patients, P_adj_ <0.05)*.

***Supplementary Figure 3: Monocyte subsets and response to ICB***

*Boxplots to show change in monocyte subset proportion with: A) cICB (n= 29 paired patient samples) and B) sICB (n= 24 paired patient samples) (Paired Kruskal Wallis test). C) Kaplan Meier curve to show lymphocyte to monocyte ratio as per flow cytometry above/below median) before treatment, C1, and overall survival (OS) (n = 57 patients) (log-rank test)*.

***Supplementary Figure 4: Single cell RNA-seq cohort details***

***Supplementary Figure 5: scRNA-seq clustering and annotation***

*A) UMAP plot to show all monocytes (from eight patients receiving ICB at baseline (C1) and day 21 post-treatment (C2), plus three healthy donor individuals (‘C’) clustered into nine subsets indicated by different colours (total cells n= 24,457) at resolution 0.25 with 13 dimensions. B) Heatmap to show proportion of assigned cell types according to SingleR annotation using expression sets including the Human Primary Cell Atlas, Monaco Immune, Immune Cell Expression and Villani datasets. SingleR was used to compare the expression of each single cell to that of an annotated expression set providing classification annotation. Only cell type annotations comprising at least 1% of the total scRNA-seq population were included. Scale (‘mat’) indicates proportion of cells in the subset assigned to that cell type with 1 indicating 100%. C) UMAP plot to show Seurat subset assignment by patient ID.. ‘C’ denotes control samples (comprises cells from three pooled healthy donor samples). Each point represents a cell and cells are coloured by Seurat cluster assignment. D) Table to show summary of annotation of Seurat monocytes clusters (Seurat clusters 0-8) with SingleR.*

*We removed subset 5 from downstream analysis as it was limited to patient ID7, and control samples. Patient ID7 was removed completely from comparisons between C1 and C2 monocyte subgroup proportion sizes*.

***Supplementary Figure 6: Characterising monocyte subsets with scRNA-seq***

*A) Heatmap to show top 20 genes by log2 fold change in classical subset comprising all Classical groups 1-4 merged (n = 21,276 cells) and CD16+ monocyte (n=840 cells) subgroups. B) Dotplot to show GOBP pathways most significantly enriched (top 15 most significant pathways, Padj<0.05) in classical and CD16+ monocyte subgroups.*

***Supplementary Figure 7: Table to show hub genes for gene expression module scoring in scRNA-seq data.***

***Supplementary Figure 8: Monocyte population responses to ICB***

*A) Boxplots to show percentage of each monocyte group per sample both before (C1) and after (C2) ICB (n=7 paired patient samples with patient ID7 removed). B) Barplot to show proportion of monocyte subgroups by condition across all patients (At C1 n= 9000 cells, at C2 n=9082 cells). C) Boxplot to show percentage per samples pre and post treatment by monocyte group for sICB patients (n=4 paired patient samples). D) Boxplot to show percentage per samples pre and post treatment by monocyte group for cICB patients (n=3 paired patient samples). E) Boxplot to show percentage of classical and CD16+ monocytes per patient sample (n=7 paired patient samples). Significance performed by a paired Wilcoxon signed rank test.*

**Table 1:** Differentially expressed genes in MM patients versus healthy donors.

**Table 2:** GOBP pathways enriched comparing MM versus healthy donors.

**Table 3:** Baseline monocyte count associated expressed transcripts

**Table 4:** monocyte count associated GOBP pathways

**Table 5:** Differentially expressed transcripts in circulating monocytes in response to ICB at day 21 (C2) across both cICB and sICB patients.

**Table 6:** cICB associated differentially expressed transcripts

**Table 7:** sICB associated differentially expressed transcripts

**Table 8:** GOBP pathways enriched across induced and suppressed transcripts following ICB at day 21 (C2).

**Table 9:** Differentially expressed transcripts preferentially induced at day 21 (C2) in those receiving cICB versus sICB.

**Table 10:** GOBP pathways enriched across transcripts preferentially induced in response to cICB versus sICB.

**Table 11:** Transcripts differentially associated with death at three months after starting ICB (C1&C2, sICB and cICB).

**Table 12:** Transcripts differentially associated with death at six months after starting ICB (C1&C2, sICB and cICB).

**Table 13:** Transcripts differentially associated with death at twelve months after starting ICB (C1&C2, sICB and cICB).

**Table 14:** GOBP pathways enriched across transcripts associated with death at three months after starting ICB (C1&C2, sICB and cICB).

**Table 15:** GOBP pathways enriched across transcripts associated with death at six months after starting ICB (C1&C2, sICB and cICB).

**Table 16:** GOBP pathways enriched across transcripts associated with death at twelve months after starting ICB (C1&C2, sICB and cICB).

**Table 17A:** Transcripts differentially associated with death at three months after starting ICB (C1 pre-treatment only, sICB and cICB).

**Table 17B:** Transcripts differentially associated with death at six months after starting ICB (C1 pre-treatment only, sICB and cICB).

**Table 17C:** Transcripts differentially associated with death at twelve months after starting ICB (C1 pre-treatment only, sICB and cICB).

**Table 18A:** Transcripts differentially associated with six month progression (C1&C2, sICB and cICB).

**Table 18B:** GOBP pathways enriched across transcripts associated with disease progression within six months of starting ICB (C1&C2, sICB and cICB).

**Table 19:** Transcripts defining monocyte subgroups (Classical1, Classical2, Classical3, Classical4, CD16+ monocytes).

**Table 20:** Transcripts defining classical monocytes versus CD16+ monocytes with scRNA-seq.

**Table 21:** GOBP pathways enriched across transcripts defining monocyte subgroups (Classical 1-4, CD16+ monocytes)

**Table 22:** GOBP pathways enriched across transcripts defining merged large classical group and CD16+ monocytes.

**Tables 23-28:** to show cohort details for scRNA-seq data and significantly correlated genes across scRNA-seq data with hub gene (pearson’s correlation, adjusted for multiple testing).

## Author contributions

BPF conceived and oversaw the study. MPF, RAW, WY, MM recruited patients. RAW annotated clinical outcome data. RAC, CT, RAW, OT, AVA, PKS, EM, SD and SK collected patient samples and isolated CD14+ cells. RAC and RAW performed the scRNA-seq experiments. CT, OT, EM and AVA performed bulk RNA-seq experiments. IN performed bioinformatic pre-processing of bulk RNA-seq and scRNA-seq data. CAT performed flow cytometry experiments. RAC performed computational analysis of the bulk RNA-seq, scRNA-seq and flow cytometry data. RAC produced figures and RAC and BPF drafted the manuscript.

## Acknowledgments and Funding

We acknowledge and thank all the patients who donated material for this study. We thank the teams at the Day Treatment Unit at the Oxford Cancer Centre for facilitating this study.

This work was funded by a Wellcome Trust Intermediate Clinical fellowship held by BPF (no. 201488/Z/16/Z), which also supported IN, SD and AVA. RAC was funded by CRUK Clinical Research Training Fellowship (S_3578). CAT was funded by the Engineering and Physical Sciences Research Council and Balliol Jowett Society (no. D4T00070). RAW was funded by a Wellcome Trust Doctoral Training Fellowship (no. BST00070). OT was funded by the Clarendon Fund, Oxford Australia Fellowship and St Edmund’s Hall. WY was supported by an NIHR ACF and a CRUK Predoctoral Fellowship (ref. RCCTI100019). MM and BPF are supported by the NIHR Oxford Biomedical Research Centre.

