## Supplementary figures and images for "Peripheral monocyte transcriptomics associated with immune checkpoint blockade outcomes in metastatic melanoma"

### Sup Fig 1

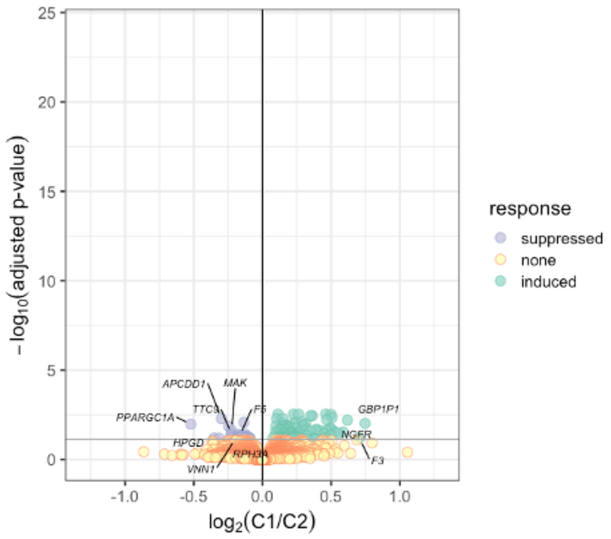


Supplementary Figure 1

### Sup Fig 2

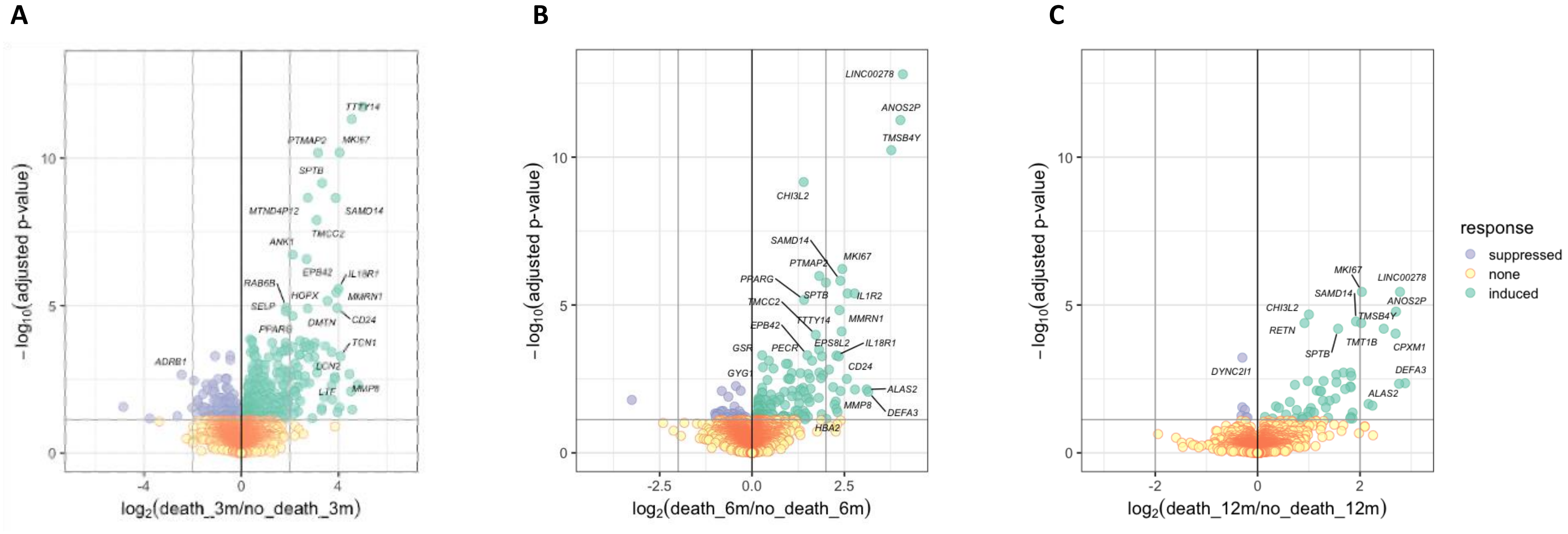


Supplementary Figure 2

### Sup Fig 3

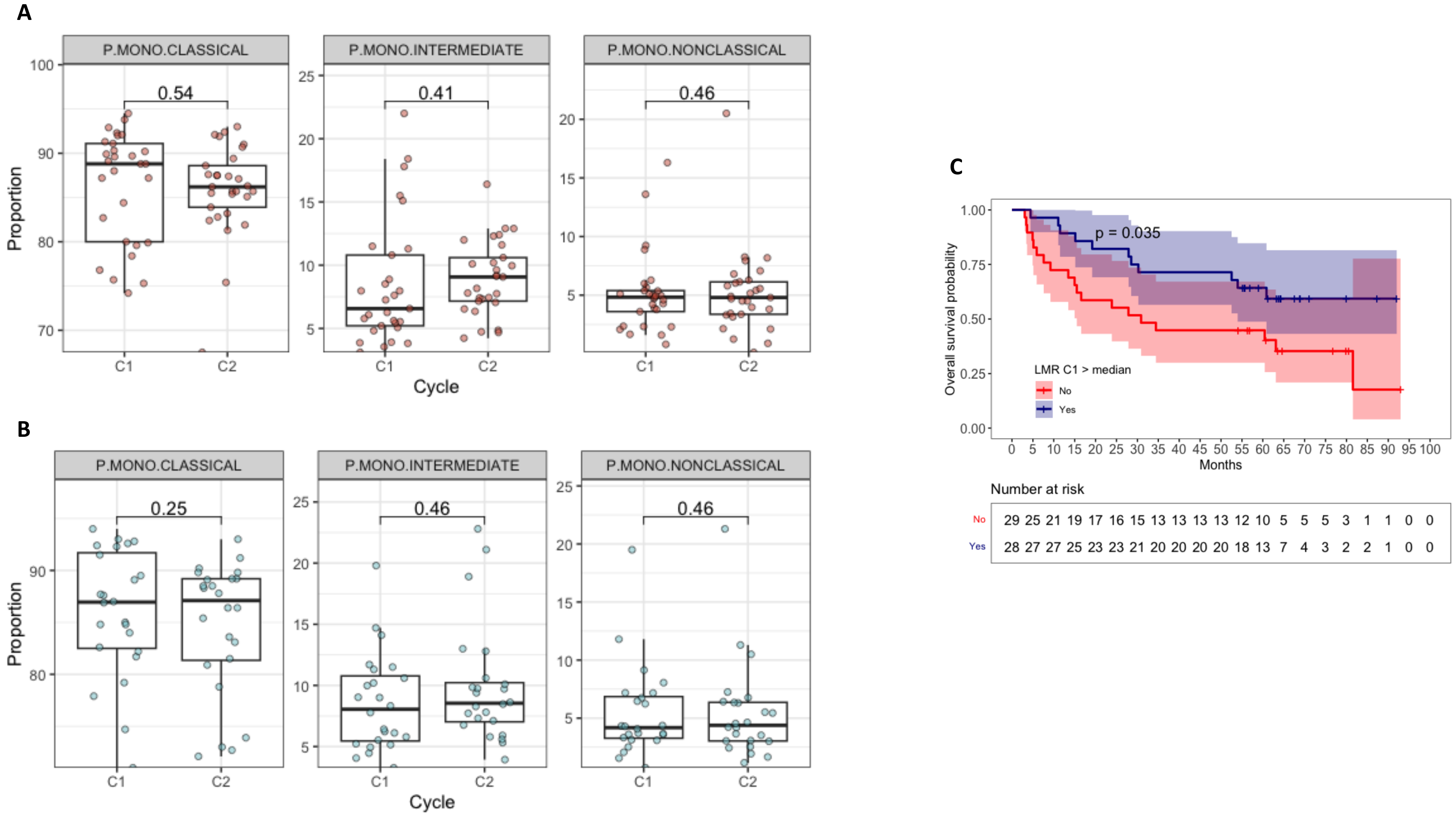


Supplementary Figure 3

### Sup Fig 6

A

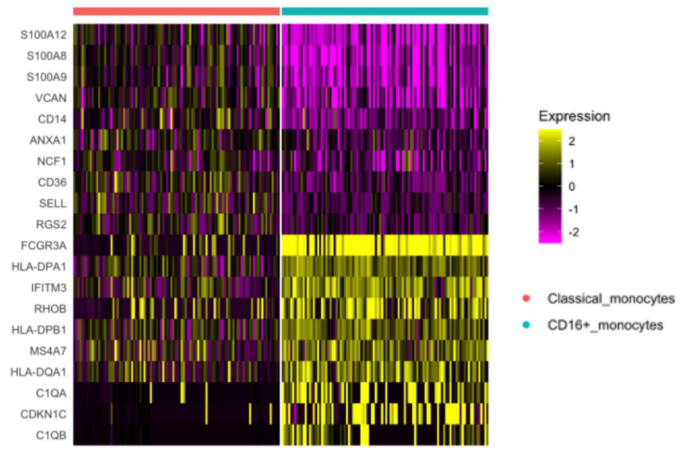


B

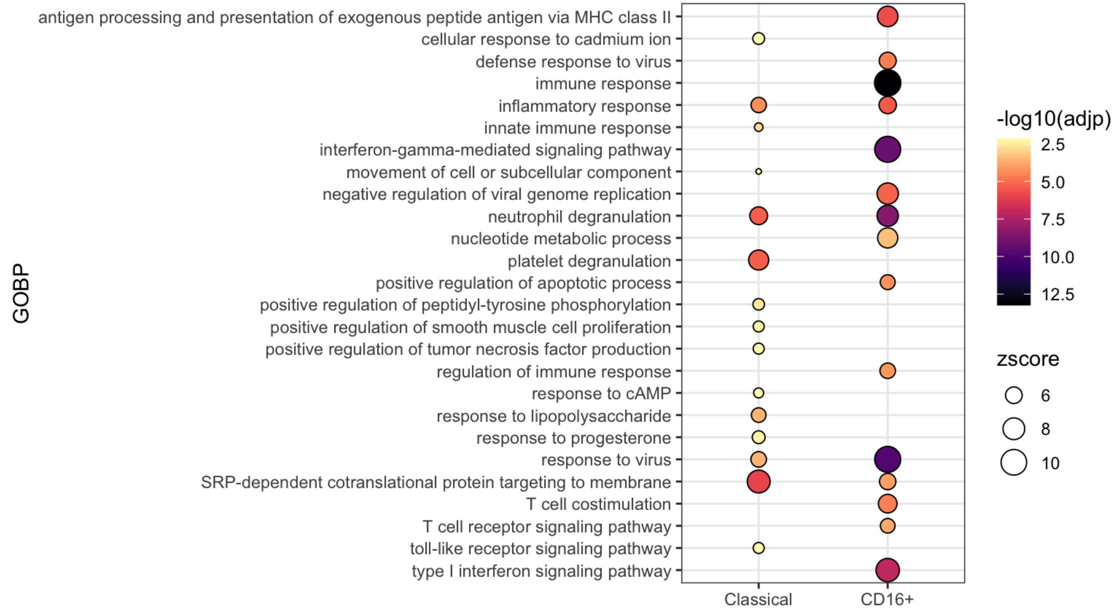


Supplementary Figure 6

### Sup Fig 8

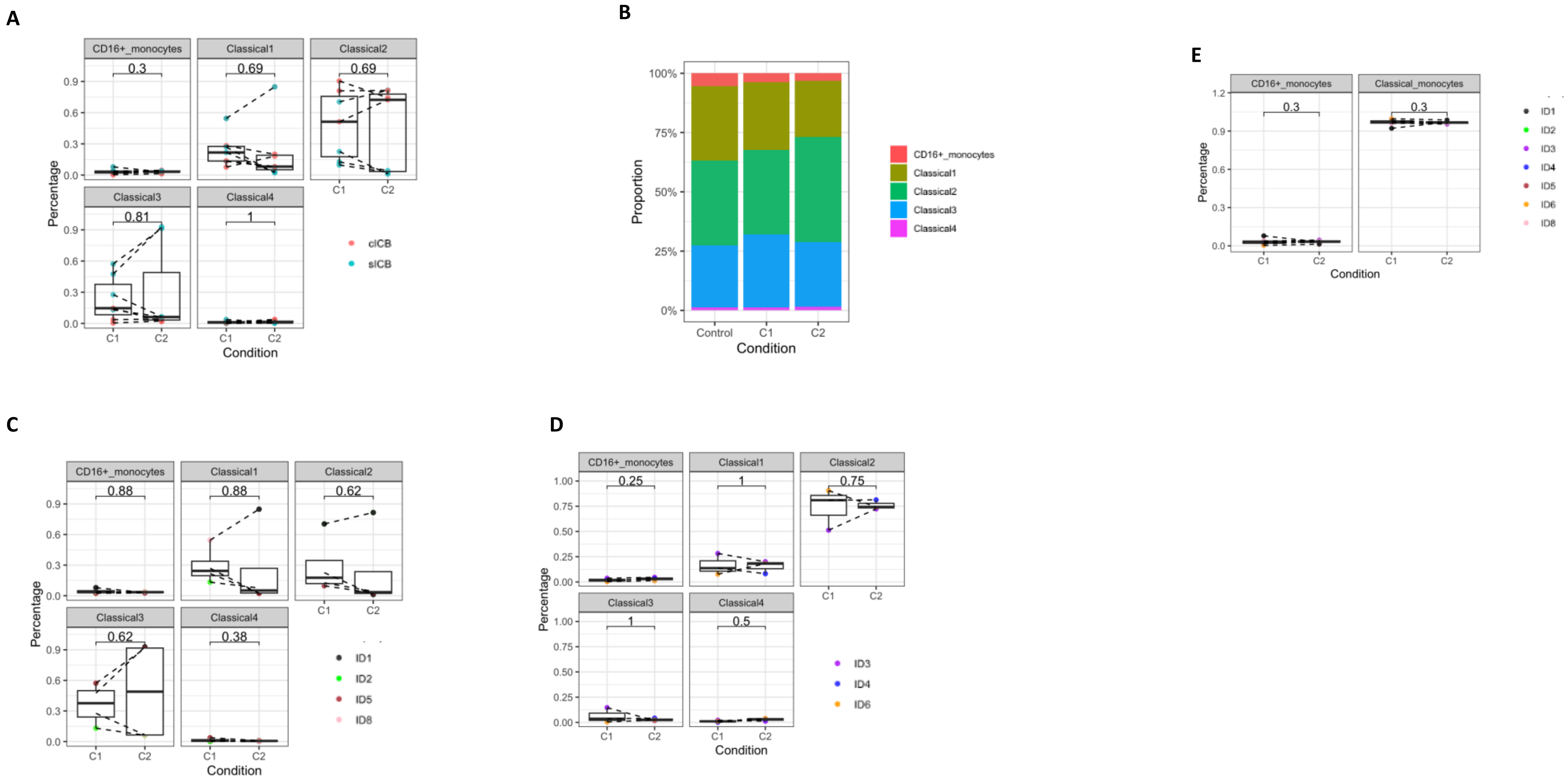


Supplementary Figure 8
