## Supplementary material for "Peripheral monocyte transcriptomics associated with immune checkpoint blockade outcomes in metastatic melanoma": Sup Fig 4

### Supplementary Figure 4

### Single cell RNA-seq Cohort details

| Patient ID | Age category | Sex | Treatment | Treatment cycle | No. $CD14^+$ cells |
|---|---|---|---|---|---|
| ID1 | 81-85 | M | sICB | C1 | 1752 |
| | | | | C2 | 1568 |
| ID2 | 71-75 | F | sICB | C1 | 475 |
| | | | | C2 | 1073 |
| ID3 | 56-60 | M | cICB | C1 | 1182 |
| | | | | C2 | 1498 |
| ID4 | 41-45 | F | cICB | C1 | 653 |
| | | | | C2 | 749 |
| ID5 | 41-45 | F | sICB | C1 | 1275 |
| | | | | C2 | 727 |
| ID6 | 56-60 | M | cICB | C1 | 695 |
| | | | | C2 | 1629 |
| ID7 | 66-70 | M | cICB | C1 | 1611 |
| | | | | C2 | 2232 |
| ID8 | 66-70 | M | sICB | C1 | 1678 |
| | | | | C2 | 1513 |
| HD_1 | 51-55 | F | NA | Control | 4147* |
| HD_2 | 56-60 | F | NA | Control | |
| HD_3 | 61-65 | F | NA | Control | |

**Appendix A: scRNA-seq cohort data**

*Table to show cohort details for scRNA-seq data. Note C1 denotes sample taken immediately pre-treatment (day1) and C2 taken day 21 post first treatment. Treatment – cICB denotes combination therapy (ipilimumab/nivolumab), whilst sICB denotes single agent therapy (pembrolizumab).*

**Control samples were pooled with library preparation, sequencing and analysis occurring on a merged Control sample from the three individuals.*
