## Supplementary material for "Peripheral monocyte transcriptomics associated with immune checkpoint blockade outcomes in metastatic melanoma": Sup Fig 5

**A**

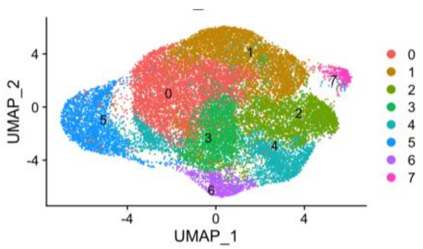


**B**

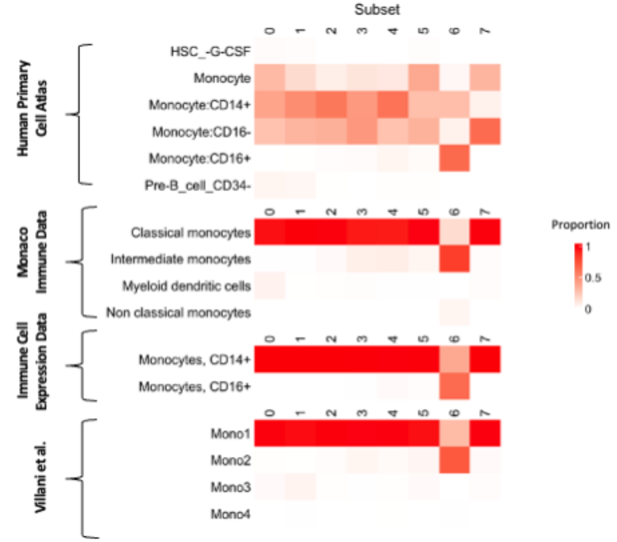


**C**

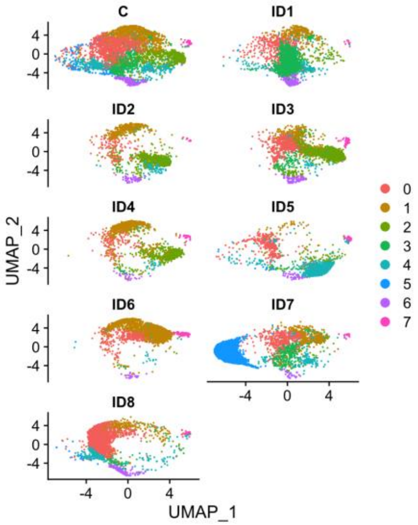


**D**

| Monocyte cluster | Annotation dataset | | | | Seurat group |
|---|---|---|---|---|---|
| | **Human Primary cell Atlas** | **Monaco Immune Data** | **Immune Cell Expression Data** | **Villani et al.** | |
| Classical | Monocyte, Monocytes CD14+, Monocyte CD16- | Classical monocytes | Monocytes, CD14+ | Mono1 | 0,1,2,3,4,5,7 |
| Intermediate | Monocyte CD16+ | Intermediate monocytes | Monocytes, CD16+ | Mono3/4 | 6 |
| Non-classical | Monocyte CD16+ | Intermediate monocytes | Monocytes, CD16+ | Mono2 | 6 |

Supplementary Figure 5
