## Supplementary material for "Peripheral monocyte transcriptomics associated with immune checkpoint blockade outcomes in metastatic melanoma": Sup Fig 7

**Supplementary Figure 7**

| Hub gene | Associated phenotype/function |
|---|---|
| CD14 | Classical monocytes |
| CCR2 | Chemotaxis |
| FCGR3A | Non-classical |
| C1QA | Complement signalling |
| STAT1 | Interferon-response |
| HLA-DRA | Antigen presentation |
